# Socioeconomic Disparities in the Effects of Pollution on Spread of Covid-19: Evidence from US Counties

**DOI:** 10.1101/2021.01.06.21249303

**Authors:** Osvaldo Allen, Ava Brown, Ersong Wang

## Abstract

This paper explores disparities in the effect of pollution on confirmed cases of Covid-19 based on counties’ socioeconomic and demographic characteristics. Using data on all US counties on a daily basis over the year 2020 and applying a rich panel data fixed effect model, we document that: 1) there are discernible social and demographic disparities in the spread of Covid-19. Blacks, low educated, and poorer people are at higher risks of being infected by the new disease. 2) The criteria pollutants including Ozone, CO, PM10, and PM2.5 have the potential to accelerate the outbreak of the novel coronavirus. 3) The disadvantaged population is more vulnerable to the effects of pollution on the spread of coronavirus. Specifically, the effects of pollution on confirmed cases become larger for blacks, low educated, and counties with lower average wages in 2019.

## 1. Introduction

The novel coronavirus was observed initially in a small cluster in Wuhan, China in December 2019 and spread around the globe during the following year, and claimed about 1.85 million deaths in 2020 (CNN, 2020). While the outbreak of the virus was unprecedented and fast the factors behind its pace of spread have risen policy-relevant questions. For instance, some studies point to the fact that there are disparities in the outbreak of the Covid-19 across occupations (McClure et al., 2020). Blacks, minorities, males, older individuals, low educated, and poorer individuals are at higher risks of being infected with Covid-19 (Figueroa et al., 2020; Kopel et al., 2020; McClure et al., 2020; Paul et al., 2020; Yang et al., 2020). For instance, Yang et al. (2020) apply a negative binomial regression at a county-level dataset that covers data on Covid-19 cases up to June 13^th^ and find that counties with a higher density of racial and ethnicity have higher confirmed cases. They show that this link is enhanced for counties with higher segregation between blacks and whites.

On the other end, there are also environmental factors that may affect the spread of Covid-19. Temperature and pollution are among the factors that were related to the Covid-19 through various mechanism channels. For instance, NoghaniBehambari, Salari, et al. (2020) examine the impacts of ambient air on the spread of Covid-19 across US counties. They use panel data fixed effect models and GMM models in a panel of county-by-day and find that an increase of one degree in air temperature is associate with 0.041 more cases per 100,000 population. The results are robust when they include county-by-week fixed effects and also across various subsamples. In another study, Contini & Costabile (2020) evaluates the literature on pollution and Covid-19 and conclude that, although marginally, specific pollutants such as PM_10_ can explain variations in the outbreak of Covid-19. However, no study has investigated the heterogeneous effects of pollution on the spread of Covid-19 based on demographic and socioeconomic characteristics. This paper aims to fill this gap in the literature.

This paper evaluates the effects of demographic and socioeconomic features of counties on the relationship between pollution and the spread of Covid-19 in the US. Using daily panel data across all US counties that cover days in the year 2020 and applying a rich panel data fixed effect model, we find that: 1) Pollution has a small but significant effect on the pace of Covid-19 outbreak. 2) There is heterogeneity in confirmed cases based on demographic characteristics. Counties with a higher share of blacks, higher share of low educated people, and lower average wages reveal higher rates of confirmed cases. 3) The marginal effects of pollution on the spread of Covid-19 is larger among counties with lower wages and a higher share of minorities.

This paper adds to the literature that investigates the sources of variations in the outbreak of Covid-19 in two ways. First, to the best of our knowledge, this is the first study to evaluate the socioeconomic disparities in the effect of pollution on Covid-19 confirmed cases. Second, we update the findings of the literature on pollution and Covid-19 using data from all US counties on a daily basis that covers all days of the year 2020 while the previous literature exploited from part of this timeframe.

The findings of this paper have important policy implications. The fact that pollution causes a mechanism channel for the spread of pandemic suggests that policymakers should re-evaluate the abatement structure during the pandemic to protect public health. The evidence on the racial and demographic disparities also help policymakers design optimal welfare programs during the pandemic to close the health gap among different groups within a society.

The rest of the paper is organized as follows. In section 2, we introduce the data sources and discuss the final sample. Section 3 provides the econometric framework. In section 4, we go over the results. Section 5 concludes the paper.

## 2. Data Sources

This study implements a wide array of data sources. the daily count of new confirmed cases is extracted from USA-Facts (2020). The daily temperature data is extracted from the Global Summary of the Day data files produced by the National Oceanic and Atmospheric Administration (NOAA). The county population and demographic data are extracted from SEER (2019). The pollution data comes from the Environmental Protection Agency (EPA). The county average wage data is from the Quarterly Census of Employment and Wages (QCEW) and is extracted from replication codes provided by NoghaniBehambari, Noghani, et al. (2020). Finally, the unemployment rate data is extracted from Local Area Unemployment Statistics gathered by the Bureau of Labor Statistics.

First, we show the geographic disparities in Covid-19 and socioeconomic disparities in a series of figures. Figure 1 illustrates the quartiles of total confirmed cases per county population (top panel) and the quartiles of daily average cases (bottom panel) across US counties for the whole year of 2020. The rates of confirmed cases for both outcomes are concentrated mainly in eastern and western states. Figure 2 depicts the geographic distribution of counties based on quartiles of the share of people with low education (top panel) and high education (bottom panel).^4^ Figure 3 shows the geographic distribution based on the percentage of whites (top panel) and the percentage of blacks (bottom panel).

**Figure 1.**
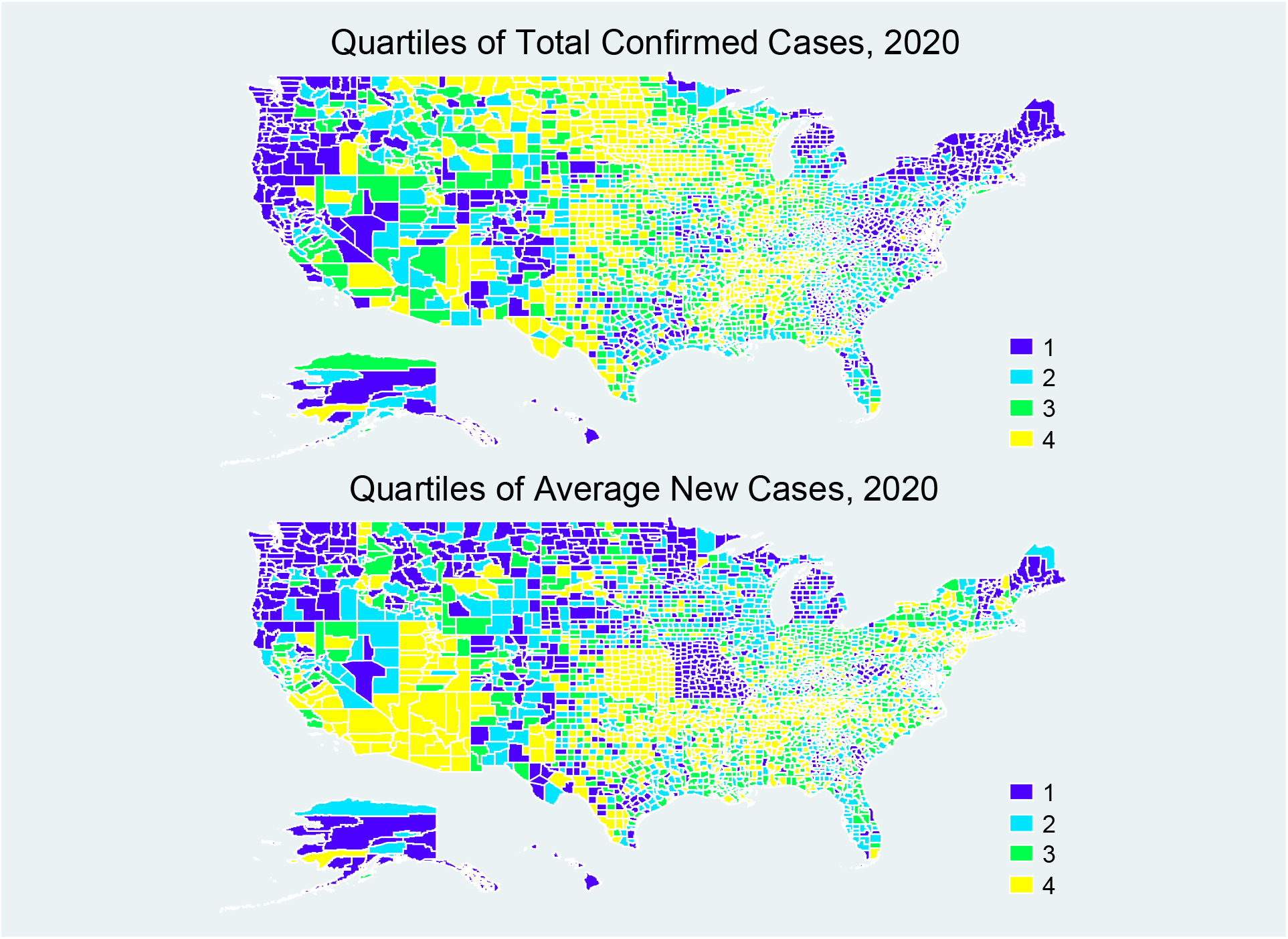
Geographic Distribution of Quartiles of Total Confirmed Cases (Top) and Average Daily Cases (Bottom) across US Counties in 2020.

**Figure 2.**
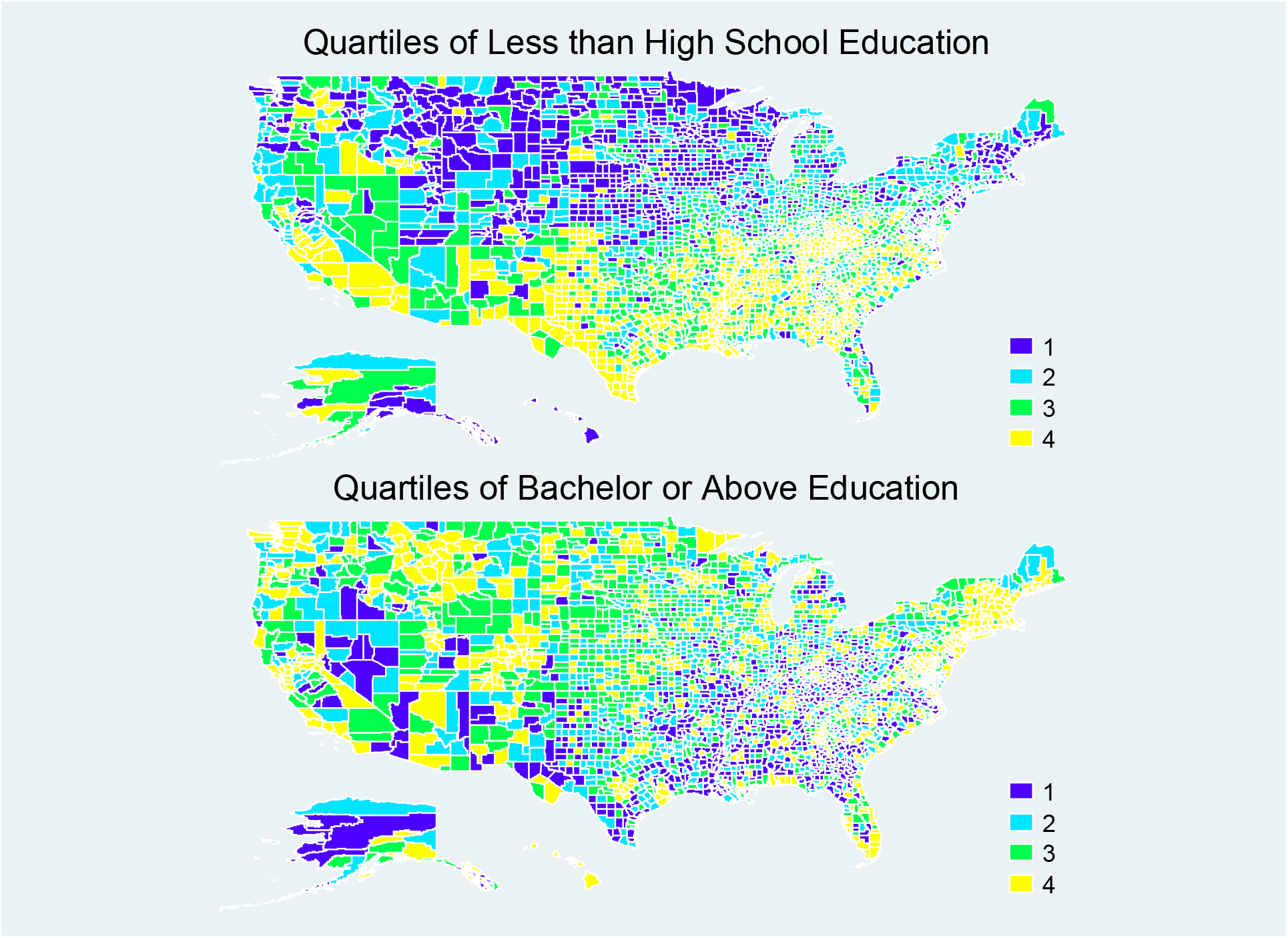
Geographic Distribution of Quartiles of Percentage of People with Less than High School Education (Top) and Bachelor and above Education (Bottom) across US Counties in 2019.

**Figure 3.**
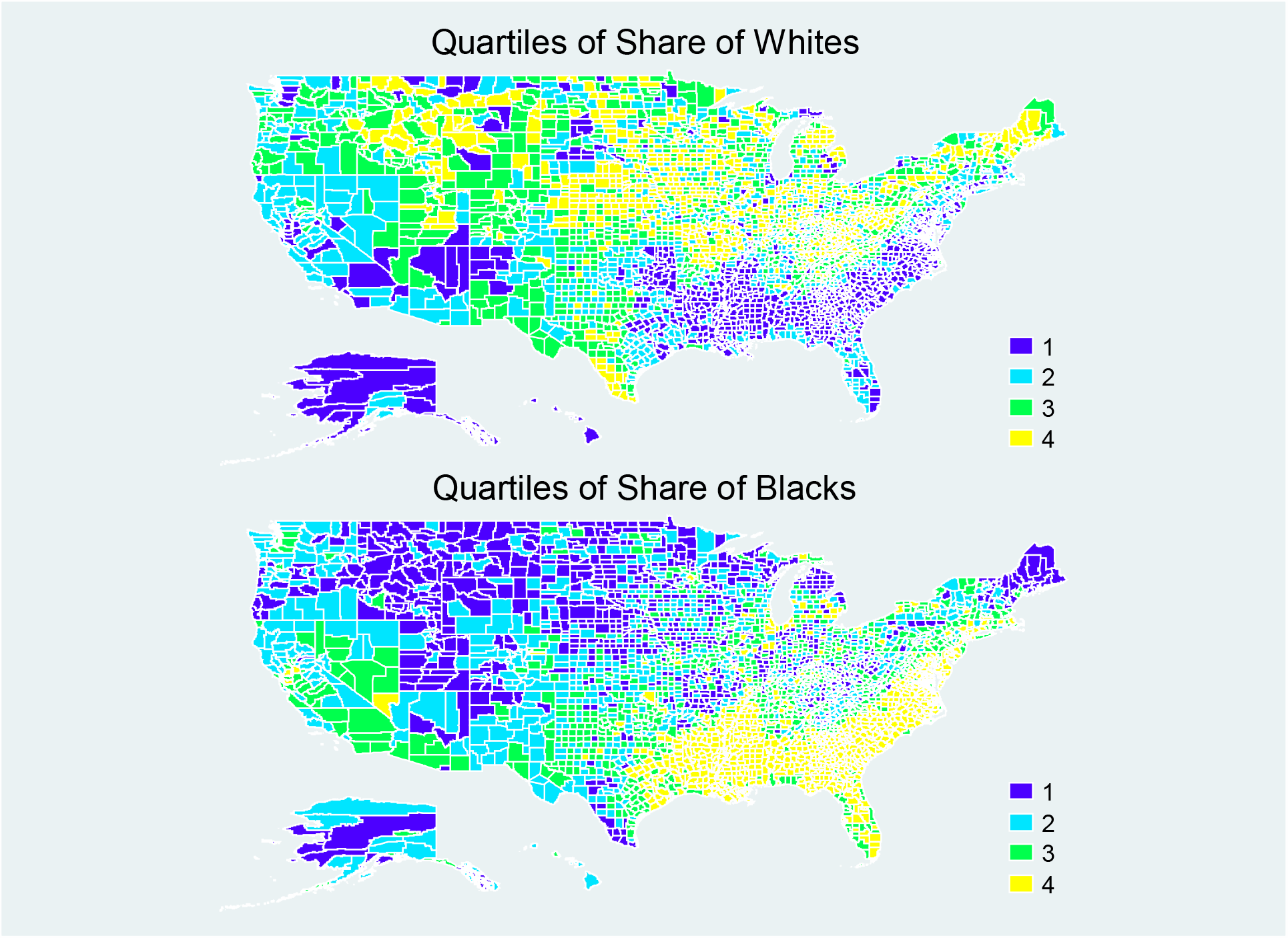
Geographic Distribution of Quartiles of Percentage of Whites (Top) and Blacks (Bottom) across US Counties in 2019.

The pollution data reported by EPA has two problems. First, pollutants have different units of measurement. In order to solve this problem and to make the interpretations easy and intuitive, we standardize the pollution data. We subtract the variable from the mean and then divide it by its standard deviation over the sample period. Therefore, all pollutants have a mean of zero and a standard deviation of one. For this reason, we avoid reporting their summary statistics. Second, the distribution of pollution monitors across counties is sporadic. Moreover, not every pollution monitor reports every essential criteria pollutant on a regular basis. To show this fact visually, Figure 4 illustrates the quartiles of Ozone pollution across counties. While the distribution is arbitrary across counties they cover a small fraction of counties. For instance, only 356 counties report Ozone among 3,148 counties covered in the final sample.

**Figure 4.**
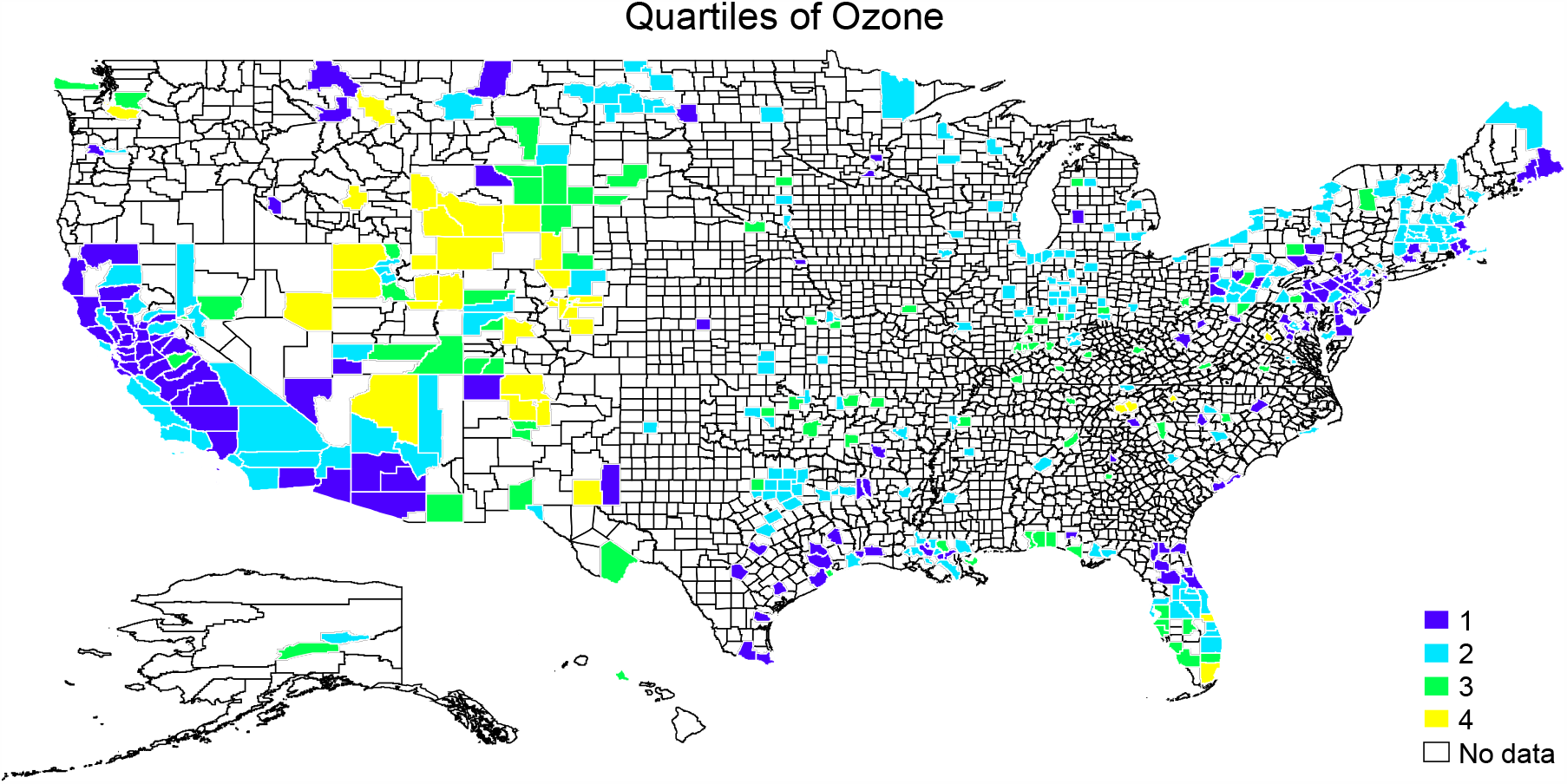
Geographic Distribution of Quartiles of Percentage of Ozone in 2020.

A summary statistics of the final sample is reported in Table 1. On average, there have been 17.8 new confirmed cases per 100,000 population and the total inflicted individuals in 2020 within each county add up to 6,588 persons per 100,000 county population. Roughly 9.9 percent of people are black and 13.4 percent are low educated.

**Table 1.**
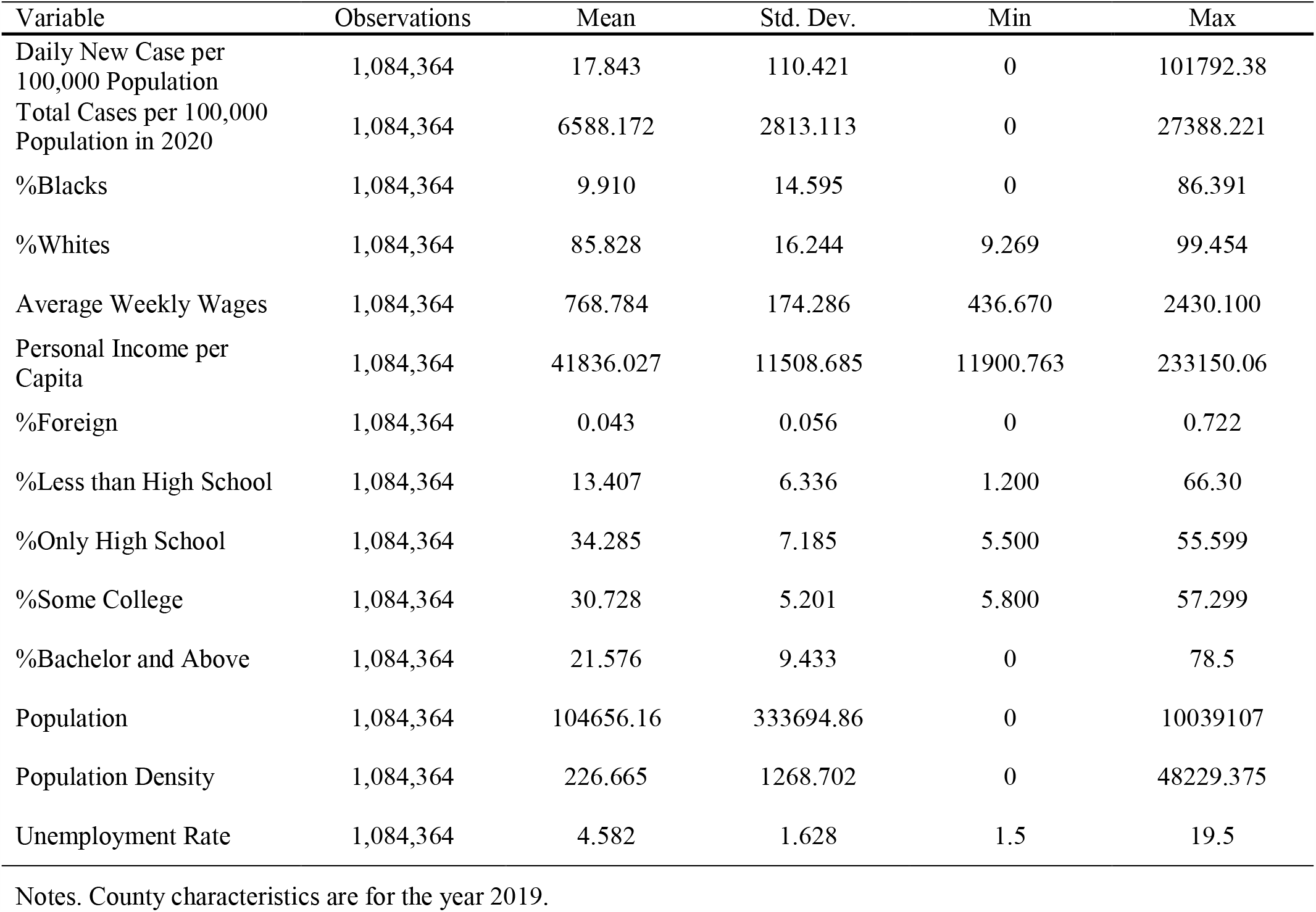
Summary Statistics.

| Variable | Observations | Mean | Std. Dev. | Min | Max |
| --- | --- | --- | --- | --- | --- |
| Daily New Case per 100,000 Population | 1,084,364 | 17.843 | 110.421 | 0 | 101792.38 |
| Total Cases per 100,000 Population in 2020 | 1,084,364 | 6588.172 | 2813.113 | 0 | 27388.221 |
| %Blacks | 1,084,364 | 9.910 | 14.595 | 0 | 86.391 |
| %Whites | 1,084,364 | 85.828 | 16.244 | 9.269 | 99.454 |
| Average Weekly Wages | 1,084,364 | 768.784 | 174.286 | 436.670 | 2430.100 |
| Personal Income per Capita | 1,084,364 | 41836.027 | 11508.685 | 11900.763 | 233150.06 |
| %Foreign | 1,084,364 | 0.043 | 0.056 | 0 | 0.722 |
| %Less than High School | 1,084,364 | 13.407 | 6.336 | 1.200 | 66.30 |
| %Only High School | 1,084,364 | 34.285 | 7.185 | 5.500 | 55.599 |
| %Some College | 1,084,364 | 30.728 | 5.201 | 5.800 | 57.299 |
| %Bachelor and Above | 1,084,364 | 21.576 | 9.433 | 0 | 78.5 |
| Population | 1,084,364 | 104656.16 | 333694.86 | 0 | 10039107 |
| Population Density | 1,084,364 | 226.665 | 1268.702 | 0 | 48229.375 |
| Unemployment Rate | 1,084,364 | 4.582 | 1.628 | 1.5 | 19.5 |

Notes. County characteristics are for the year 2019.

## 3. Econometric Framework

We start with a cross-sectional data of counties and explore the cross-tabulation between county characteristics in 2019 and the rate of spread of Covid-19 in 2020 using the following OLS model:

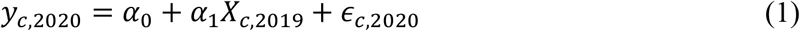

The main reason to use the characteristics in 2019 is that socioeconomic characteristics have not yet been released for the year 2020. In this specification, *y* is the Covid-19 confirmed cases per 100,000 population of county *c* for the year 2020.

In the next step, we use a panel of county-by-day data to assess the effect of pollution on the rates of Covid-19 using fixed-effect models of the following form:

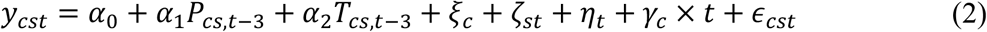

Where *c* Indexes the county, *s* indexes the state, and *t* indexes day-by-month of observation. *y* is the daily rates of Covid-19 confirmed in the county. *P* is the standardized variable of pollution measures including Ozone, Carmon Monoxide (CO), particulate matters less than 10*μm* (PM_10_), and particulate matters less than 2.5*μm* (PM_2.5_). The pollution is measured three days in advance since the literature suggests that the virus has an average incubation period of 3 days (Lauer et al., 2020; Li et al., 2020; Tan et al., 2005). Since temperature is discussed to be one of the causes that accelerate the outbreak we also control for daily temperature in all regressions represented by *T* (NoghaniBehambari, Salari, et al., 2020; Wang et al., 2020; Xie & Zhu, 2020). The parameter *ξ* is the county fixed effect. *ζ* is a set of state by day-month fixed effects. The matrix *η* represents day-by-month fixed effects. In some specifications, we also include a county-specific linear time trend. Finally, *ϵ* is a disturbance term. All standard errors are clustered at the county level. All regressions are weighted using the average of county population in 2020. To assess the socioeconomic disparities in *α*_1_ of equation 2, we use an interaction term for each characteristics using the following formulation:

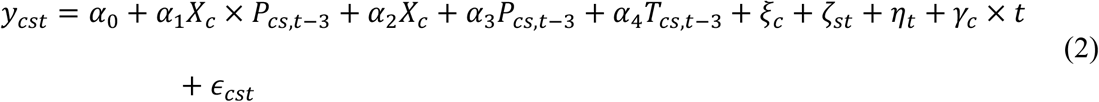

Where all parameters follow the same notation as in equations 1 and 2. The coefficient of interest s *α*_1_ that shows the effect of pollution on coronavirus cases per population for the group with characteristics represented in *X* compared to the reference group.

## 4. Main Results

We start by reevaluating the social disparities in confirmed cases of coronavirus. Table 2 shows the results of regressions introduced in equation 1 for total and average daily cases in columns 1 and 2, respectively. If the share of blacks in a county goes up by 10 percent the confirmed cases of corona increase by 1.1 cases per 100,000 population, an increase equivalent to a 6.2 percent change from the mean of daily confirmed cases (column 2, first row). In a similar manner, if the share of people with at least a bachelor’s degree goes up by 10 percent in a county then the average daily confirmed cases go down by 16.9 cases per 100,000 population. This change can explain 15.5 percent of the standard deviation of confirmed cases over the year 2020. Overall, counties with a higher share of blacks, low educated people, and lower-income and wages have higher rates of confirmed cases.

**Table 2.** Socioeconomic Disparities in the Spread of Covid-19 across US Counties.

|  | Total Case Rate in<br>2020 | Average Daily<br>Cases |
| --- | --- | --- |
|  | (1) | (2) |
| %Blacks | 2.500***<br>(0.891) | 0.011***<br>(0.0024) |
| $R^2$ | 0.001 | 0.001 |
| %Whites | -3.931***<br>(0.691) | -0.007***<br>(0.001) |
| $R^2$ | 0.001 | 0.001 |
| Average Weekly<br>Wages | -1.783***<br>(0.232) | -0.0048***<br>(0.0006) |
| $R^2$ | 0.012 | 0.009 |
| Per Capita Personal<br>Income (\$1,000) | -23.091***<br>(5.277) | 0.063***<br>(0.011) |
| $R^2$ | 0.009 | 0.006 |
| %Less than High<br>School | 63.507***<br>(8.201) | 0.171***<br>(0.022) |
| $R^2$ | 0.020 | 0.014 |
| %Some College | 26.004**<br>(10.371) | 0.072**<br>(0.029) |
| $R^2$ | 0.002 | 0.002 |
| %Bachelor and<br>Above | -59.221***<br>(4.703) | -0.169***<br>(0.014) |
| $R^2$ | 0.039 | 0.031 |
| Observations | 3,135 | 3,135 |

Notes. Each cell represents a separate regression. Robust standard errors are reported in parentheses.

Next, we reexamine the effect of pollution on Covid-19. Using equation 2, Table 3 reports the results for models without and with a linear county trend (columns 1 and 2, respectively). Each independent variable is in a separate row and each cell represents a separate regression. Looking at the full specification of column 2, one standard deviation increase in CO, Ozone, PM_10_, and PM_2.5_ is associated with an increase in Covid-19 cases by 0.04, 0.29, 0.35, and 0.11 cases per 100,000 population. Although these effects are marginal and economically small they are significant at 1% level and robust to including or excluding county by time linear trend.

**Table 3.**
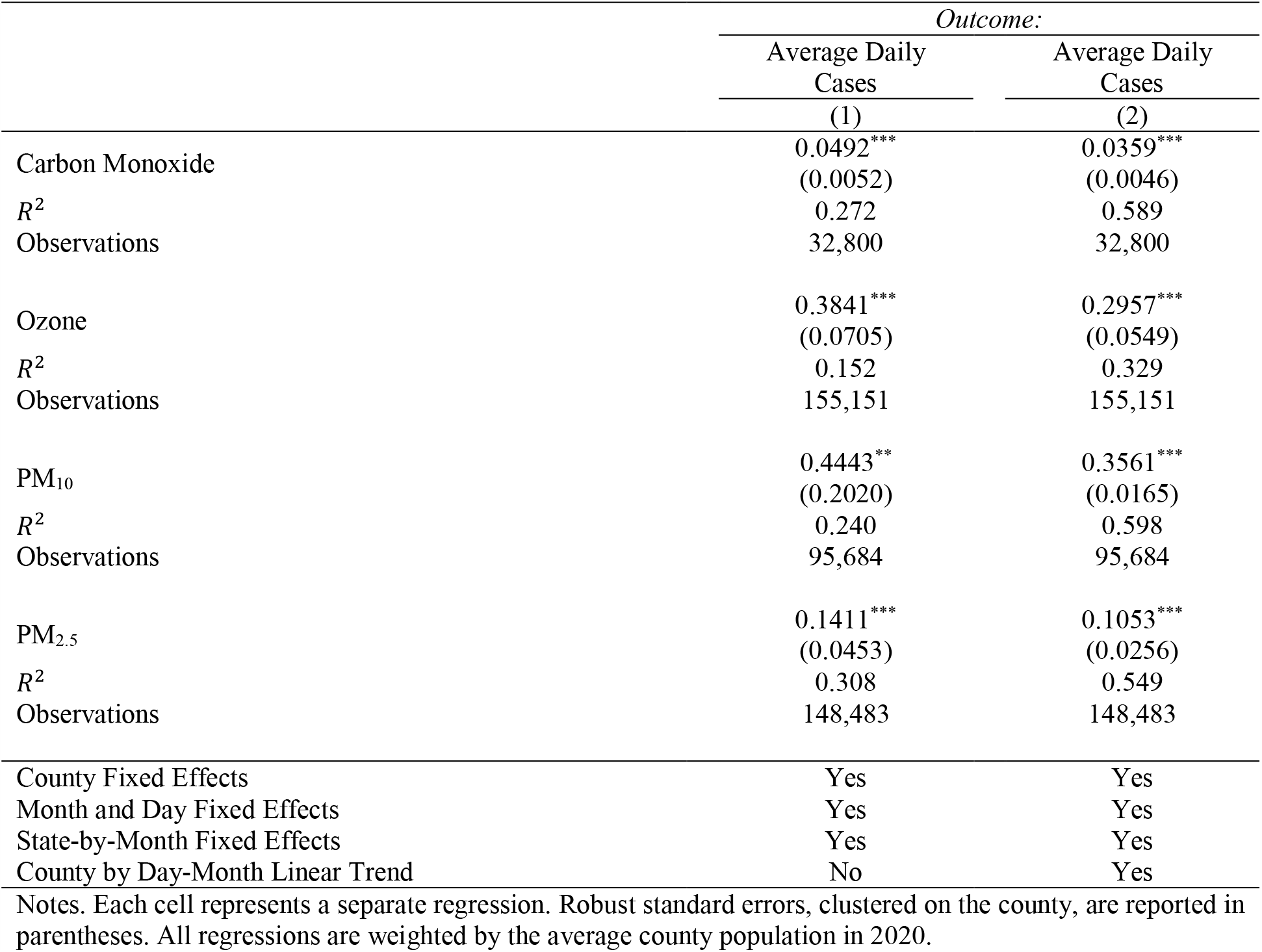
The Effect of Criteria Pollutants on Spread of Covid-19.

Finally, we report the main results of the paper using equation 3 in Table 4 through Table 7. Interestingly, as areas with a higher share of poor people and minorities reveal higher confirmed cases they also are more susceptible to pollution-driven confirmed cases. The interaction term between pollution measures and blacks (Table 4) and low educated (Table 6) are positive implying that the relationship between pollution and the outbreak of the virus is stronger among these people. On the other hand, the interaction term between pollutants and whites (Table 5), high educated (Table 7), and average wages (Table 8) are negative implying the protective effects against the Covid-19 consequence of pollution among counties with a higher share of whites, high educated, and income. For instance, the marginal effect of one standard deviation increase in PM_10_ on confirmed cases for a 10 percent rise in the share of blacks in a county goes up by 6.49 cases per 100,000 population (column 3, Table 4).

**Table 4.** The Heterogeneity of the Effects of Criteria Pollutants on Spread of Covid-19 among Blacks.

|  | *Outcome: Average Daily Cases* |  |  |  |
| --- | --- | --- | --- | --- |
|  | (1) | (2) | (3) | (4) |
| Carbon Monoxide × %Blacks | 0.095***<br>(0.015) | | | |
| Ozone × %Blacks | | 0.518***<br>(0.095) | | |
| PM<sub>10</sub> × %Blacks | | | 0.649***<br>(0.026) | |
| PM<sub>2.5</sub> × %Blacks | | | | 0.238***<br>(0.046) |
| County Fixed Effects | Yes | Yes | Yes | Yes |
| Month and Day Fixed Effects | Yes | Yes | Yes | Yes |
| State-by-Month Fixed Effects | Yes | Yes | Yes | Yes |
| County by Day-Month Linear Trend | Yes | Yes | Yes | Yes |
| $R^2$ | 0.59 | 0.33 | 0.59 | 0.57 |
| Observations | 32,800 | 155,151 | 95,684 | 148,483 |

Notes. Each cell represents a separate regression. Robust standard errors, clustered on the county, are reported in parentheses. All regressions are weighted by the average county population in 2020.

**Table 5.** The Heterogeneity of the Effects of Criteria Pollutants on Spread of Covid-19 among Whites.

|  | *Outcome: Average Daily Cases* |  |  |  |
| --- | --- | --- | --- | --- |
|  | (1) | (2) | (3) | (4) |
| Carbon Monoxide × %Whites | -0.026***<br>(0.003) | | | |
| Ozone × %Whites | | -0.221***<br>(0.061) | | |
| PM<sub>10</sub> × %Whites | | | -0.298***<br>(0.049) | |
| PM<sub>2.5</sub> × %Whites | | | | -0.092***<br>(0.012) |
| County Fixed Effects | Yes | Yes | Yes | Yes |
| Month and Day Fixed Effects | Yes | Yes | Yes | Yes |
| State-by-Month Fixed Effects | Yes | Yes | Yes | Yes |
| County by Day-Month Linear Trend | Yes | Yes | Yes | Yes |
| $R^2$ | 0.59 | 0.33 | 0.59 | 0.57 |
| Observations | 32,800 | 155,151 | 95,684 | 148,483 |

Notes. Each cell represents a separate regression. Robust standard errors, clustered on the county, are reported in parentheses. All regressions are weighted by the average county population in 2020.

**Table 6.**
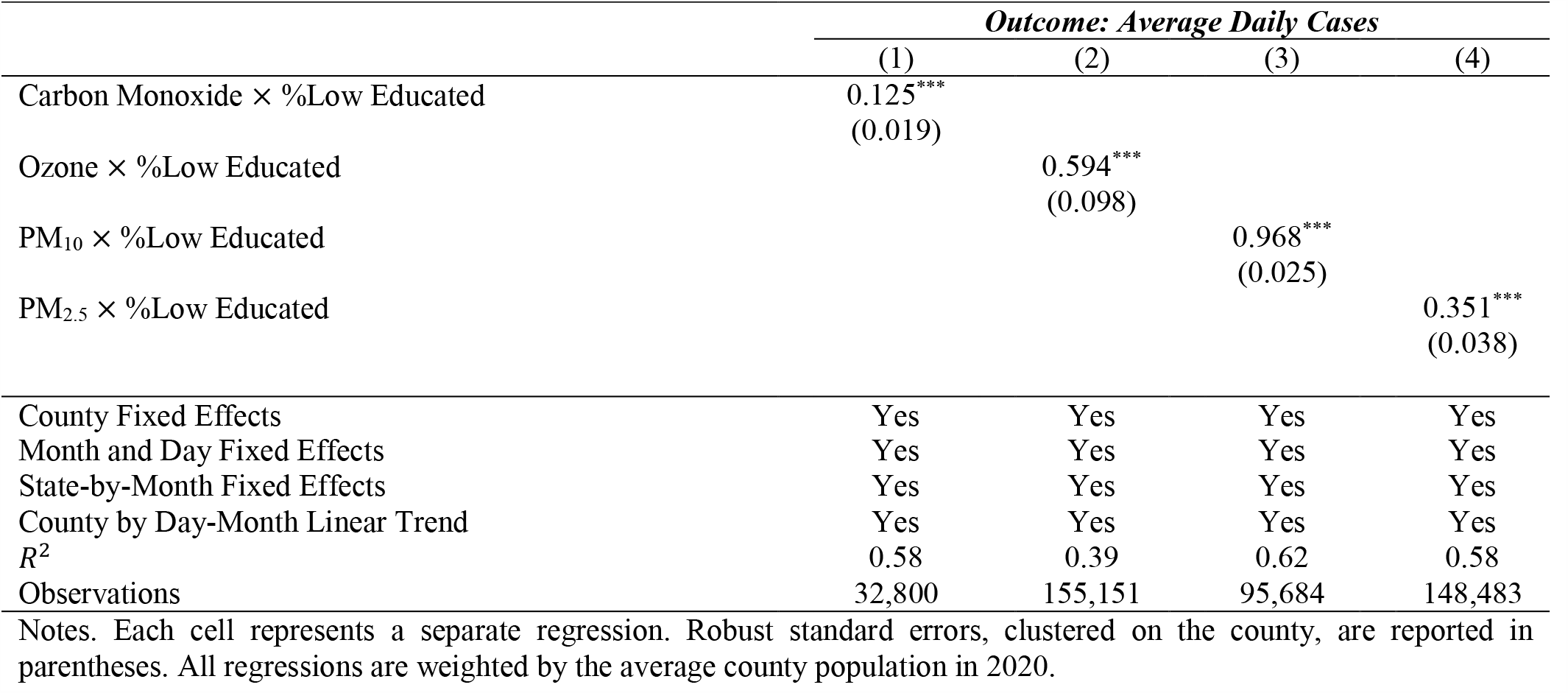
The Heterogeneity of the Effects of Criteria Pollutants on Spread of Covid-19 among Low Educated.

**Table 7.** The Heterogeneity of the Effects of Criteria Pollutants on Spread of Covid-19 among High Educated.

|  | *Outcome: Average Daily Cases* |  |  |  |
| --- | --- | --- | --- | --- |
|  | (1) | (2) | (3) | (4) |
| Carbon Monoxide × %High Educated | -0.015***<br>(0.003) | | | |
| Ozone × %High Educated | | -0.087**<br>(0.041) | | |
| PM<sub>10</sub> × %High Educated | | | -0.124*<br>(0.087) | |
| PM<sub>2.5</sub> × %High Educated | | | | -0.087***<br>(0.014) |
| County Fixed Effects | Yes | Yes | Yes | Yes |
| Month and Day Fixed Effects | Yes | Yes | Yes | Yes |
| State-by-Month Fixed Effects | Yes | Yes | Yes | Yes |
| County by Day-Month Linear Trend | Yes | Yes | Yes | Yes |
| $R^2$ | 0.58 | 0.39 | 0.62 | 0.58 |
| Observations | 32,800 | 155,151 | 95,684 | 148,483 |

Notes. Each cell represents a separate regression. Robust standard errors, clustered on the county, are reported in parentheses. All regressions are weighted by the average county population in 2020.

**Table 8.** The Heterogeneity of the Effects of Criteria Pollutants on Spread of Covid-19 based on Average Wages.

|  | *Outcome: Average Daily Cases* |  |  |  |
| --- | --- | --- | --- | --- |
|  | (1) | (2) | (3) | (4) |
| Carbon Monoxide × Average Wages | -0.029***<br>(0.003) | | | |
| Ozone × Average Wages | | -0.265***<br>(0.046) | | |
| PM<sub>10</sub> × Average Wages | | | -0.342***<br>(0.016) | |
| PM<sub>2.5</sub> × Average Wages | | | | -0.098***<br>(0.023) |
| County Fixed Effects | Yes | Yes | Yes | Yes |
| Month and Day Fixed Effects | Yes | Yes | Yes | Yes |
| State-by-Month Fixed Effects | Yes | Yes | Yes | Yes |
| County by Day-Month Linear Trend | Yes | Yes | Yes | Yes |
| $R^2$ | 0.63 | 0.45 | 0.71 | 0.67 |
| Observations | 32,800 | 155,151 | 95,684 | 148,483 |

Notes. Each cell represents a separate regression. Robust standard errors, clustered on the county, are reported in parentheses. All regressions are weighted by the average county population in 2020.

## 5. Conclusion

Understanding the racial and social disparities in exposure to a pandemic and specifically, the disparities in the effect of pollution on the outbreak of a pandemic are essential for policymakers to design optimal welfare programs and effective restriction orders. In this paper, we explored this aspect of the outbreak of Covid-19 using daily data across all US counties covering all days of 2020. Applying a rich set of fixed effects that also controls for a linear county by time trend, we documented that 1) there are discernible social and demographic disparities in the spread of Covid-19. Blacks, low educated, and poorer people are at higher risks of being infected by the new disease. 2) The criteria pollutants have the potential to accelerate the outbreak of the virus. Among others, these pollutants include Ozone, CO, PM_10_, and PM_2.5_. 3) The disadvantaged population is more vulnerable to the effects of pollution on the spread of coronavirus. Specifically, the effects of pollution on confirmed cases become larger for blacks, low educated, and counties with lower average wages in 2019. Overall, these results suggest that the abatement structures should be strengthened during a pandemic with more weight towards areas with a higher concentration of minorities and poor people.

## Data Availability

The data is available upon request.

## Footnotes

4 Throughout the paper we categorize people with less than high school education as low educated. Similarly, we consider people with bachelor and above as high educated.

